# Predicting elevated natriuretic peptide in chest radiography: Emerging utilization gap for artificial intelligence

**DOI:** 10.1101/2023.02.22.23286205

**Authors:** Eisuke Kagawa, Masaya Kato, Noboru Oda, Eiji Kunita, Michiaki Nagai, Aya Yamane, Shogo Matsui, Yuki Yoshitomi, Hiroto Shimajiri, Tatsuya Hirokawa, Shunsuke Ishida, Genki Kurimoto, Keigo Dote

**Affiliations:** Department of Cardiology, Hiroshima City Asa Hospital, Hiroshima, Japan; Department of Cardiology, Hiroshima City North Medical Center Asa Citizens Hospital, Hiroshima, Japan

**Keywords:** Deep learning, Neural network, Machine learning, Brain natriuretic peptide, Heart failure

## Abstract

**Aims:** This study assessed an artificial intelligence (AI) model’s performance in predicting elevated brain natriuretic peptide (BNP) levels from chest radiograms and its effect on human diagnostic performance.

**Methods and results:** Patients who underwent chest radiography and BNP testing on the same day were included. Data were sourced from two hospitals: one for model development, and the other for external testing. Two final ensemble models were developed to predict elevated BNP levels of >= 200 pg/mL and >= 100 pg/mL, respectively. Humans were evaluated to predict elevated BNP levels, followed by the same test, referring to the AI model’s predictions. The 8390 images from 1334 patients were collected for model creation, and 1713 images from 273 patients for tests. The AI model achieved an accuracy of 0.855, precision of 0.873, sensitivity of 0.827, specificity of 0.882, f1 score of 0.850, and receiver-operating-characteristics area-under-curve of 0.929. The accuracy of the testing with the 100 images by 35 participants significantly improved from 0.708±0.049 to 0.829±0.069 (P < 0.001) with the AI assistance (an accuracy of 0.920). Without the AI assistance, the accuracy of the experts was higher than that of non-experts (0.728±0.051 vs. 0.692±0.042, P = 0.030); however, with the AI assistance, the accuracy of the non-experts was rather higher than that of the experts (0.851±0.074 vs. 0.803±0.054, P = 0.033).

**Conclusion:** The AI model can predict elevated BNP levels from chest radiograms and has the potential to improve human performance. The gap in utilizing new tools represents one of the emerging issues.

**Graphical Abstract:** We developed AI models using an ensemble method to predict elevated BNP levels. The AI model achieved a higher accuracy rate than any individual participant. While the accuracy of experts was higher in the non-assisted test, with the AI assistance, the accuracy of non-experts surpassed that of the experts. AI, artificial intelligence; AUC, area-under-curve; BNP, brain natriuretic peptide; GPU, graphic processing unit; PR, precision-recall; ROC, receiver-operating-characteristics.

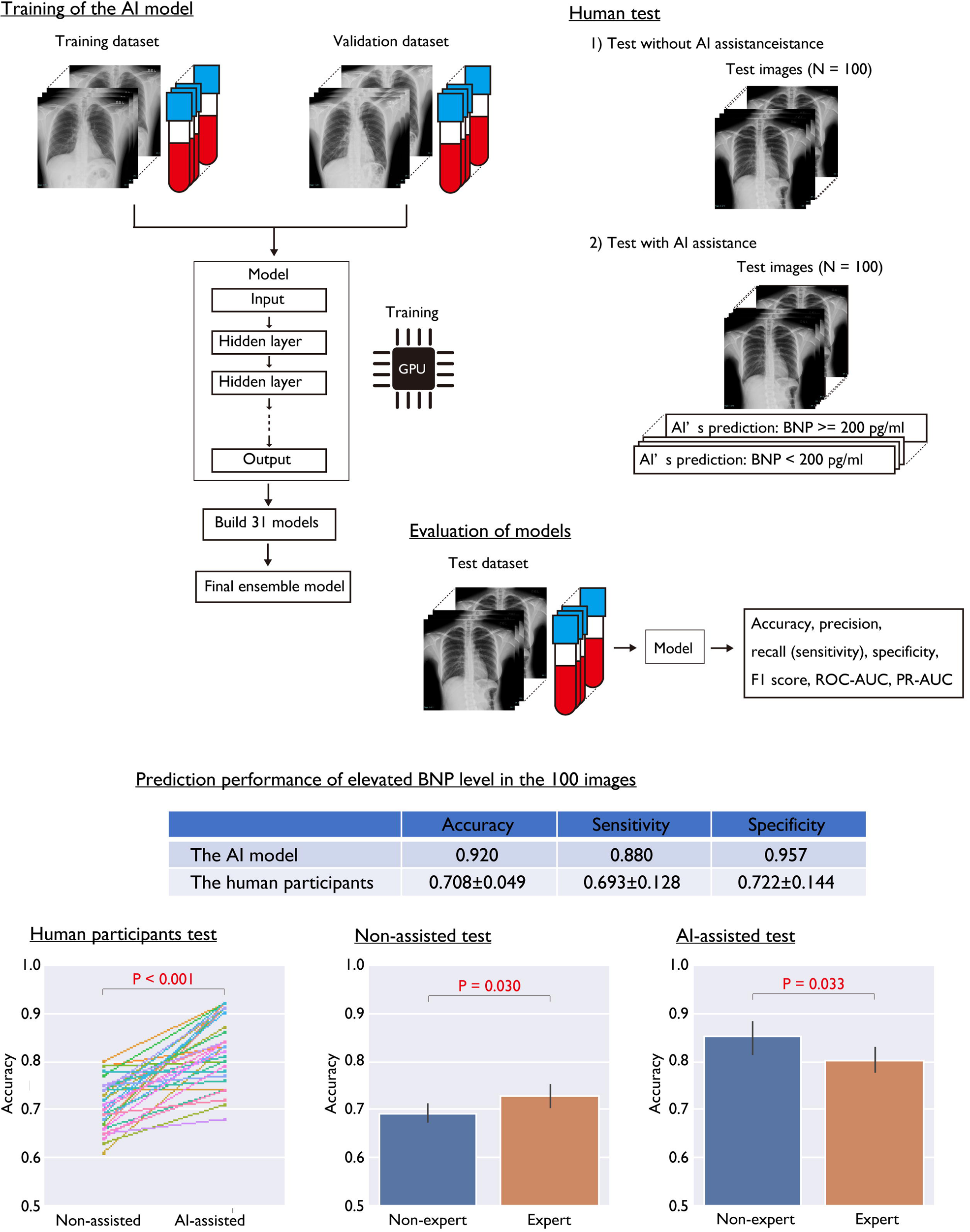

## Introduction

Heart failure is a major cause of visits to medical facilities in both unplanned emergency situations and routine medical checkups. It is also a growing and called heart failure pandemic because of the aging population.^1^ However, diagnosing heart failure, which should be medically managed, can be challenging due to the variability of symptoms, especially in heart failure that is complicated by other diseases for patients visiting medical facilities or for attending physicians who are not familiar with cardiovascular disease.^2^ Several tests are used to evaluate heart failure. Chest X-rays are widely available, and the images can be obtained quickly; however, the evaluation of chest X-ray images requires experience and has limited sensitivity and specificity for heart failure.^3^ Natriuretic peptide levels are useful not only for diagnosing heart failure but also for heart failure management.^4–6^ However, natriuretic peptide testing requires equipment, and even if the facility has such equipment, it may not be available at all hours, as many facilities do not offer testing at night or on weekends. Furthermore, it is essential to make the decision to perform the test itself. We hope that automated support tools will be developed to assist in our daily practice of heart failure quickly and inexpensively. The rise of artificial intelligence (AI) and the evolution of computer hardware provide novel findings and solutions.^7,8^ With regard to image recognition, deep neural networks (deep learning) provide relatively good performance compared to those of previous architectures, and some have already been deployed in clinical practice.^9–11^ The aim of our study was to diagnose heart failure using chest X-ray imaging and an AI model, and to support clinical practice. We hypothesized that AI models that predict elevated brain natriuretic peptide (BNP) levels from chest X-ray images could provide excellent performance compared to experienced cardiologists and could improve the diagnostic performance of humans.

## Methods

### Study Patients and Datasets

Patients who underwent chest radiography and BNP testing on the same day at Hiroshima City Asa Hospital and Hiroshima City North Medical Center Asa Citizens Hospital from October 2021 to September 2022 were eligible for this study. Since BNP testing is the first choice for natriuretic peptide testing in these hospitals, and not N-terminal pro-brain natriuretic peptide, we selected BNP for this study. We reviewed the medical records of eligible patients, including periods other than the above, and when chest X-ray and BNP testing were performed on the same day, the chest X-ray images, and plasma BNP values were collected. To increase the robustness and generalizability of the AI models, all conditions of the frontal view of chest X-ray images were collected, including anterior-posterior, posterior-anterior, standing, sitting, and supine positions, with or without inspiration, and any diseases or conditions. Lateral chest radiographs were not included in this study. According to the statement of the Japanese Heart Failure Society, we used a BNP cut-off value of 200 pg/mL for the main study and 100 pg/mL for the sub study.^12^ The chest X-ray images were assigned a binary label according to the cut-off value. The patients from Hiroshima City Asa Hospital were used for the training and validation datasets, while the patients from Hiroshima City North Medical Center Asa Citizens Hospital were used for the external test dataset. The study patients from Hiroshima City Asa Hospital were randomly divided into two datasets for training and validation. The patients were assigned to datasets in the training: validation: testing dataset ratio of approximately 0.66:0.17:0.17. Many patients had multiple pairs of chest X-ray images and BNP labels, and each patient’s data were assigned to only one dataset to avoid overfitting. The models used in this study, along with the sample code for their utilization, will be made available on GitHub following the publication of this study. The study was approved by the local institutional review board.

### Outline of an AI Model

We fine-tuned 31 modified pre-trained image recognition models as weak learners to predict elevated BNP levels, and subsequently created an ensemble model. Details are provided in the Supplemental Methods. The Proposed Requirements for Cardiovascular Imaging-Related

### Evaluation of Models

After obtaining the 31 models (weak learners), we constructed the final soft ensemble model by averaging the probabilities of the 31 models. Probabilities >= 0.5 were considered to represent BNP levels >= the cut-off value. The accuracy, precision, sensitivity (recall), specificity, F1 score, receiver-operating-characteristics (ROC) curves, and precision-recall (PR) curves were calculated using the test dataset. ROC and PR curves were constructed using probability of BNP >= cut-off value, and the area under the curve (AUC) was calculated. To avoid overfitting the test dataset, the results of the performance tests using the test dataset were not used to retrospectively train or select the models.

### Human Testing

We evaluated human performance to predict elevated BNP levels from chest radiography. The subjects were voluntary participants from the hospitals’ staff. The general findings of heart failure seen in chest X-ray images, as well as the characteristics of BNP, were taught to those being evaluated. The test subjects were shown chest X-ray images and their corresponding BNP labels in the training dataset for their learning phase. Then, they evaluated the 100 chest X-ray images from the test dataset and provided their binary prediction. The 100 images for human testing comprised 50 images with BNP < the cut-off value and 50 with BNP <= the cut-off value, presented in random order. After the first test, to assess whether the AI assistance could improve human diagnostic performance, the test subjects evaluated the same 100 images again, this time with reference to the predictions of the AI model. The performance of the AI model, which had the accuracy of 86% on the test dataset (approximately 10 to 20% higher than that of humans), was explained before the second test. The accuracy of the AI model for the 100 images and the ratio of the two labels were not disclosed to the test subjects until all tests were completed. An expert was defined as someone with a medical career of >= 10 years.

### Statistical Analysis and Calculations

Continuous variables are presented as medians (with first and third quartiles) or as mean ± standard deviation (SD), and categorical variables are presented as numbers and percentages, as appropriate. We utilized Python 3.10.7 (Python Software Foundation, Delaware, USA) and TensorFlow 2.10.1 (Google LCC, Mountain View, CA, USA) for our machine learning and statistical analysis. The difference in the accuracies of the first and second human tests was tested using Welch’s t-test or paired t-test, as appropriate. A P value < 0.05 was considered statistically significant.

## Results

### Baseline Characteristics

An overview of this study is shown in Graphical Abstract, and the baseline characteristics of the study patients are shown in Table 1. No data were missing. Among the 1607 patients in the study, the diagnoses included heart failure (N = 471), acute heart failure (N = 320), coronary artery disease (N = 517), acute coronary syndrome (N = 176), hypertrophic cardiomyopathy (N = 32), interstitial pneumonia (N = 64), and hemodialysis (N = 23). In total, 10103 chest X-ray images were collected. These images were divided among the training, validation, and test datasets as follow: 1061 patients (66%) with 6697 images (66%), 273 patients (17%) with 1693 images (17%), and 273 patients (17%) with 1713 images (17%).

**Table 1.** Characteristics of the Study Patients and Materials.

| Patient |  |
| --- | --- |
| Study patients | 1607 |
| Male sex | 980 (61) |
| Diagnosis (multiple)<sup>a</sup> |  |
| Heart failure | 471 (29) |
| Acute heart failure | 320 (20) |
| Coronary artery disease | 517 (32) |
| Acute coronary syndrome | 176 (11) |
| Hypertrophic cardiomyopathy | 32 (2) |
| Congenital heart disease | 14 (1) |
| Atrial fibrillation | 253 (16) |
| Peripheral artery disease | 97 (6) |
| Pericardial effusion | 6 (1) |
| Aortic disease | 68 (4) |
| Hemodialysis | 23 (1) |
| Chronic kidney disease | 171 (11) |
| Pneumonia | 62 (4) |
| Interstitial pneumonia | 64 (4) |
| Venous thromboembolism | 43 (3) |
| Pneumothorax | 9 (1) |
| Lung carcinoma | 16 (1) |
| Trauma | 5 (1) |
| Chest pain syndrome | 29 (2) |
| Dataset, patients |  |
| Train | 1061 (66) |
| Valid | 273 (17) |
| Test | 273 (17) |
| Images |  |
| Chest X-ray images, N | 10103 |
| Age, y | 74 (64 – 81) |
| Plasma BNP, pg/mL | 158 (47 – 569) |
| Dataset, images |  |
| Train | 6697 (66) |
| BNP ≥ 200 pg/mL | 2994 (45) |
| Valid | 1693 (17) |
| BNP ≥ 200 pg/mL | 726 (43) |
| Test | 1713 (17) |
| BNP ≥ 200 pg/mL | 852 (50) |

Data presented as N (%) or median (interquartile range). BNP, brain natriuretic peptide. <sup>a</sup>Multiple selections were allowed to account for patients with co-existing conditions.

### Performance of the AI

The 31 models (weak learners) were created and trained (Figure 1). Their performance is detailed in Table 2, Graphical Abstract, Figure 2, and Figure 3. The performance metrics of the final ensemble model were as follows: accuracy was 0.855, precision was 0.873, sensitivity (recall) was 0.827, specificity was 0.882, F1 score was 0.850, ROC AUC was 0.929, and PR AUC was 0.934.

**Figure 1.**
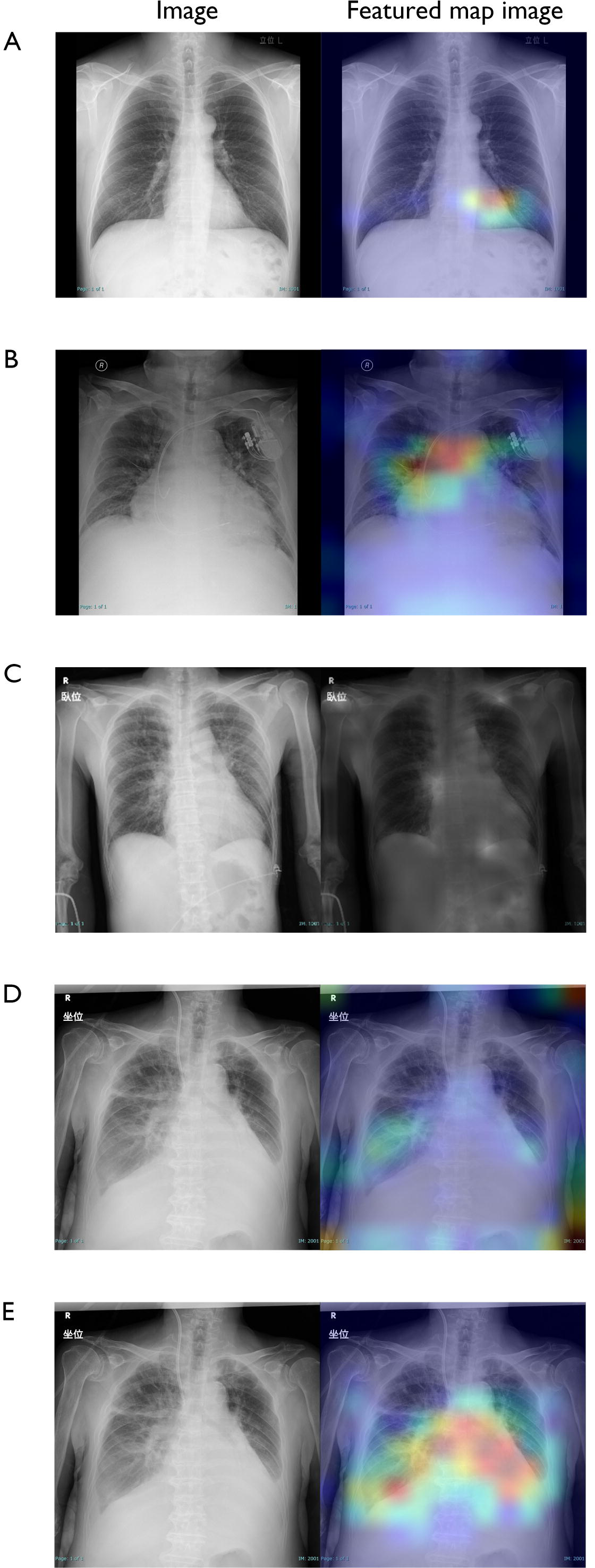
Chest X-Ray Images and Featured Map of Weak Learners. (A) Chest X-ray image with a BNP of 9 pg/mL and a GRAD-CAM image generated by the EfficientNetV2L-based-model. (B) The EfficientNetV2L-based-model identified features such as pulmonary congestion and pacemaker, with a BNP of 798 pg/mL. (C) The attention map revealed the Vit-b16-based-model’s features in the caption, the pulmonary vessel, and the diaphragmatic line, with a BNP of 413 pg/mL. The caption in the upper left corner is a Kanji character meaning the supine position. (D) The EfficientNetV2S-based-model identified features in the captions of images with a BNP of 824 pg/mL. The caption in the upper left corner is a Kanji character meaning the sitting position. (E) The other EfficientnetV2S-based-model did not focus on the features in the captions but on the pleural effusion, cardiomegaly, pulmonary artery, air bronchogram, and Kerley A lines. The caption in the upper left corner is a Kanji character meaning the sitting position. BNP, brain natriuretic peptide; GRAD-CAM, gradient-class activation maps.

**Figure 2.**
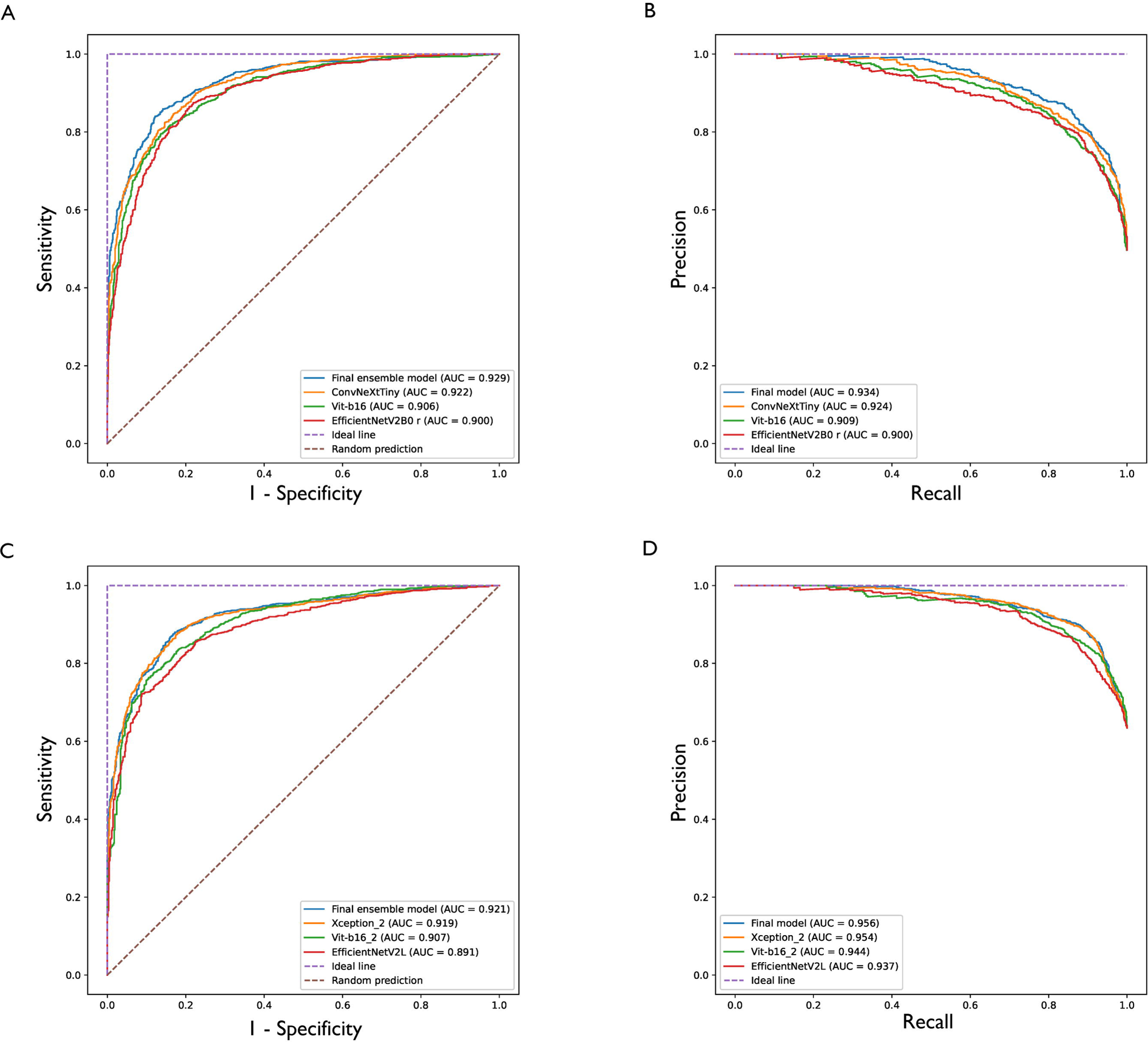
Performance of the Models. The ROC curves (A) and PR curves (B) of the AI models for predicting BNP >= 200 pg/mL are shown. Similarly, the ROC curves (C) and PR curves (D) for predicting BNP >= 100 pg/mL are shown. The ROC and PR curves were shown for the final ensemble model as well as for 3 of the 31 weak leaners. BNP, brain natriuretic peptide; PR, precision-recall; ROC, receiver-operating-characteristics.

**Figure 3.**
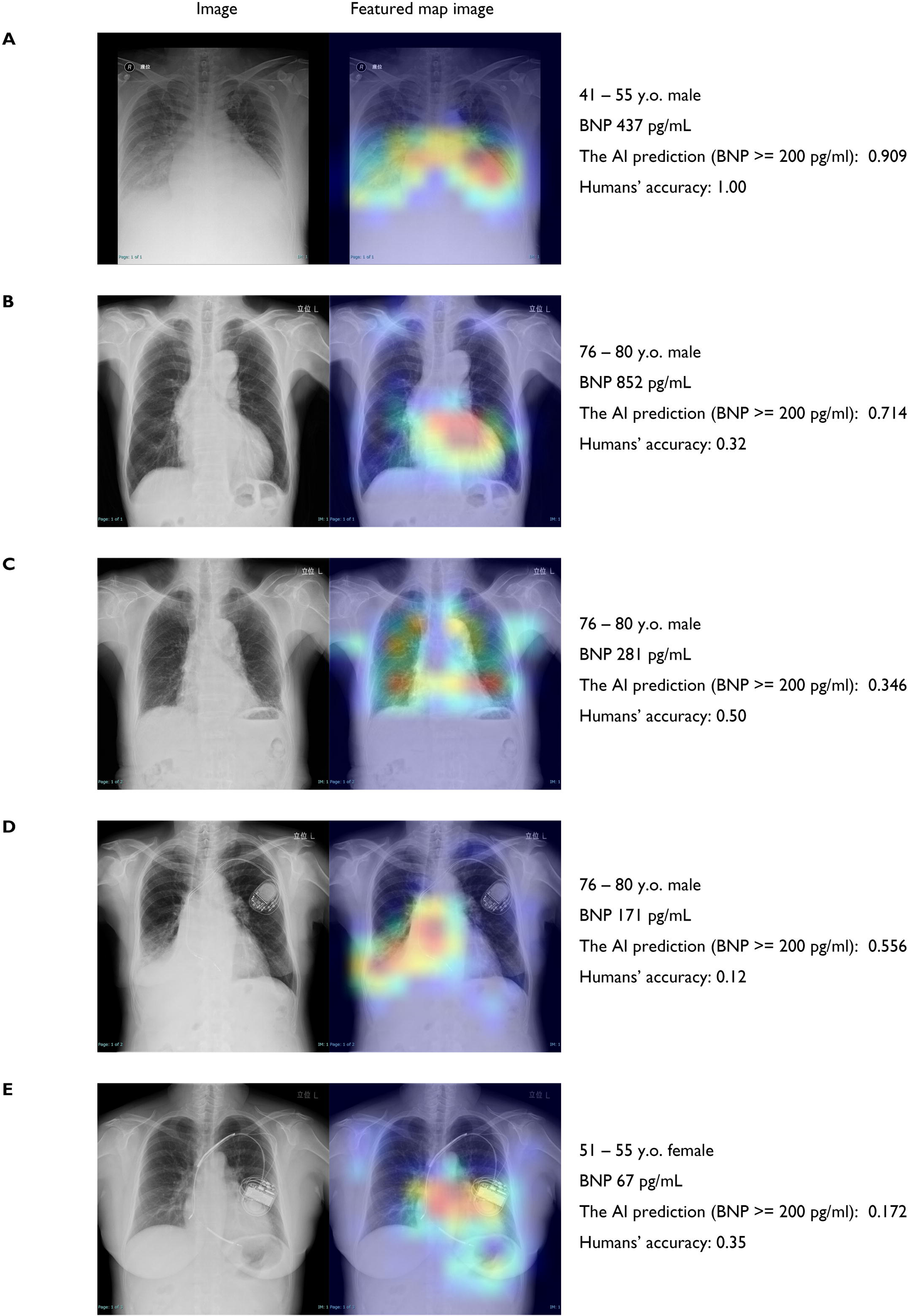
Representative Images and Predictions of the AI Models and Humans. The chest X-ray images and their featured map images, age, sex, BNP value, AI prediction, and human accuracy are shown. The featured map images were generated by the EfficientNetV2S-based-model using GRAD-CAM. The caption in the upper right corner of (A) and the upper left corner of (B, C, and D) are Kanji character meaning sitting and standing position, respectively. AI, artificial intelligence; BNP, brain natriuretic peptide; GRAD-CAM, gradient-class activation maps.

**Table 2.**
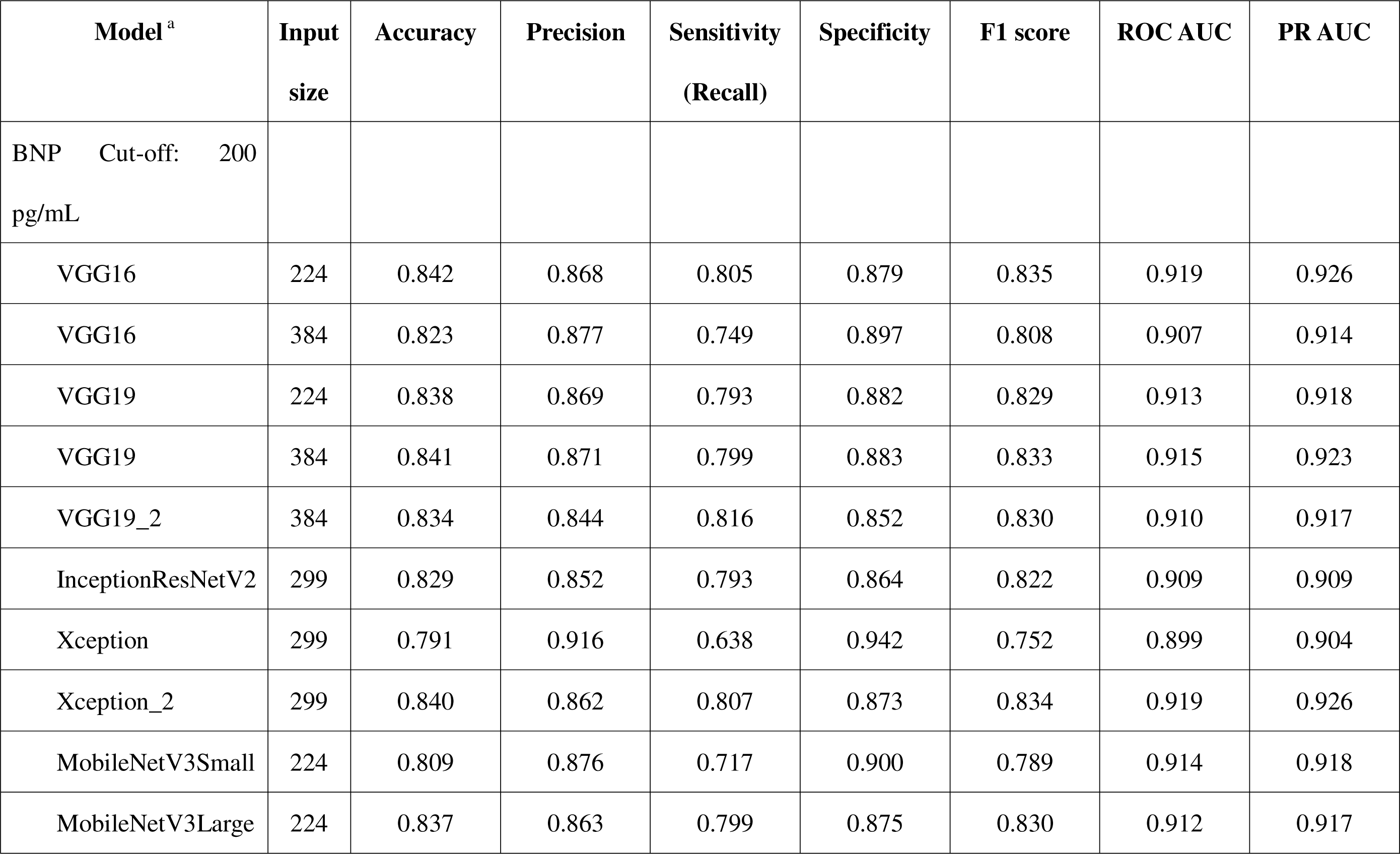
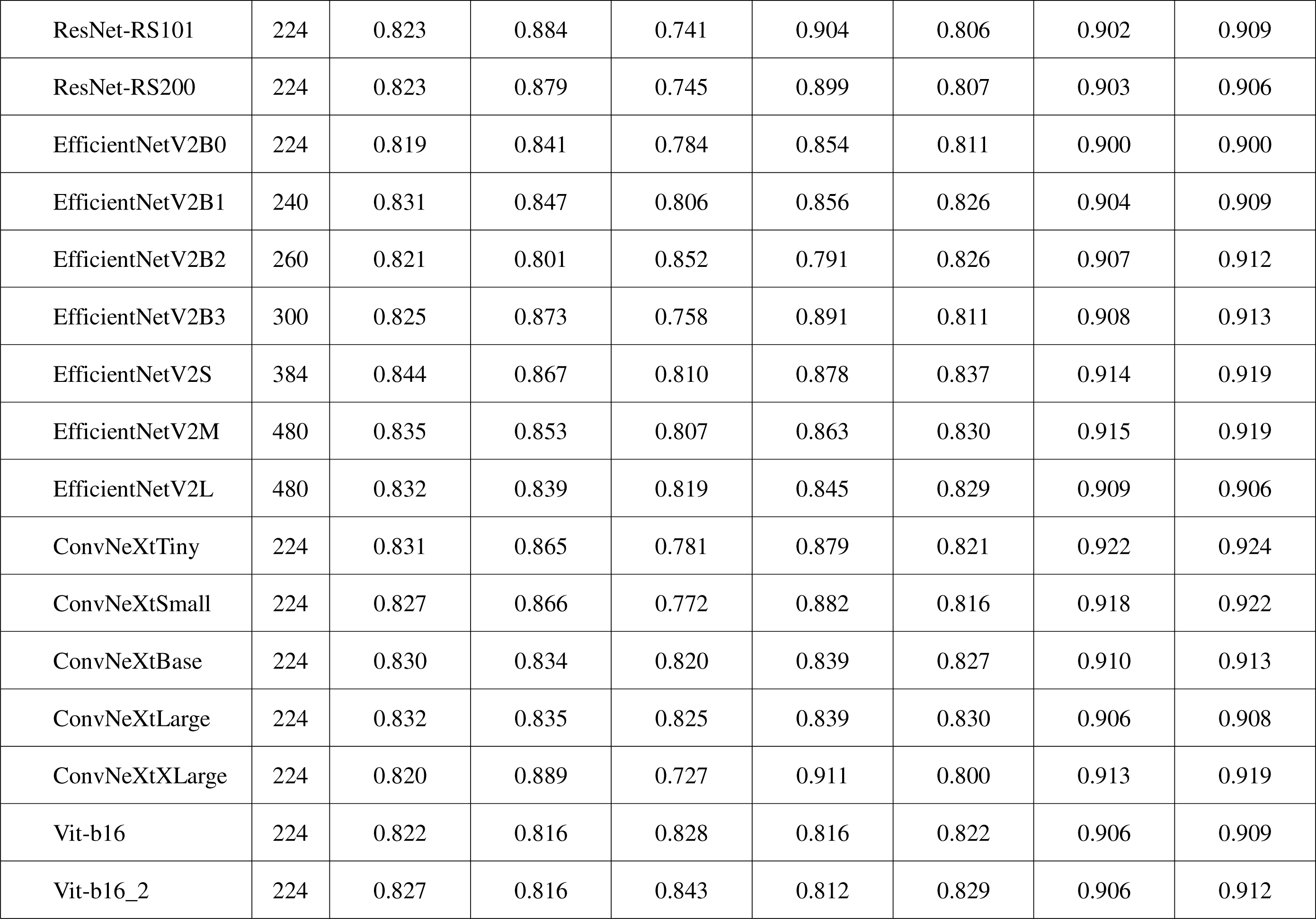
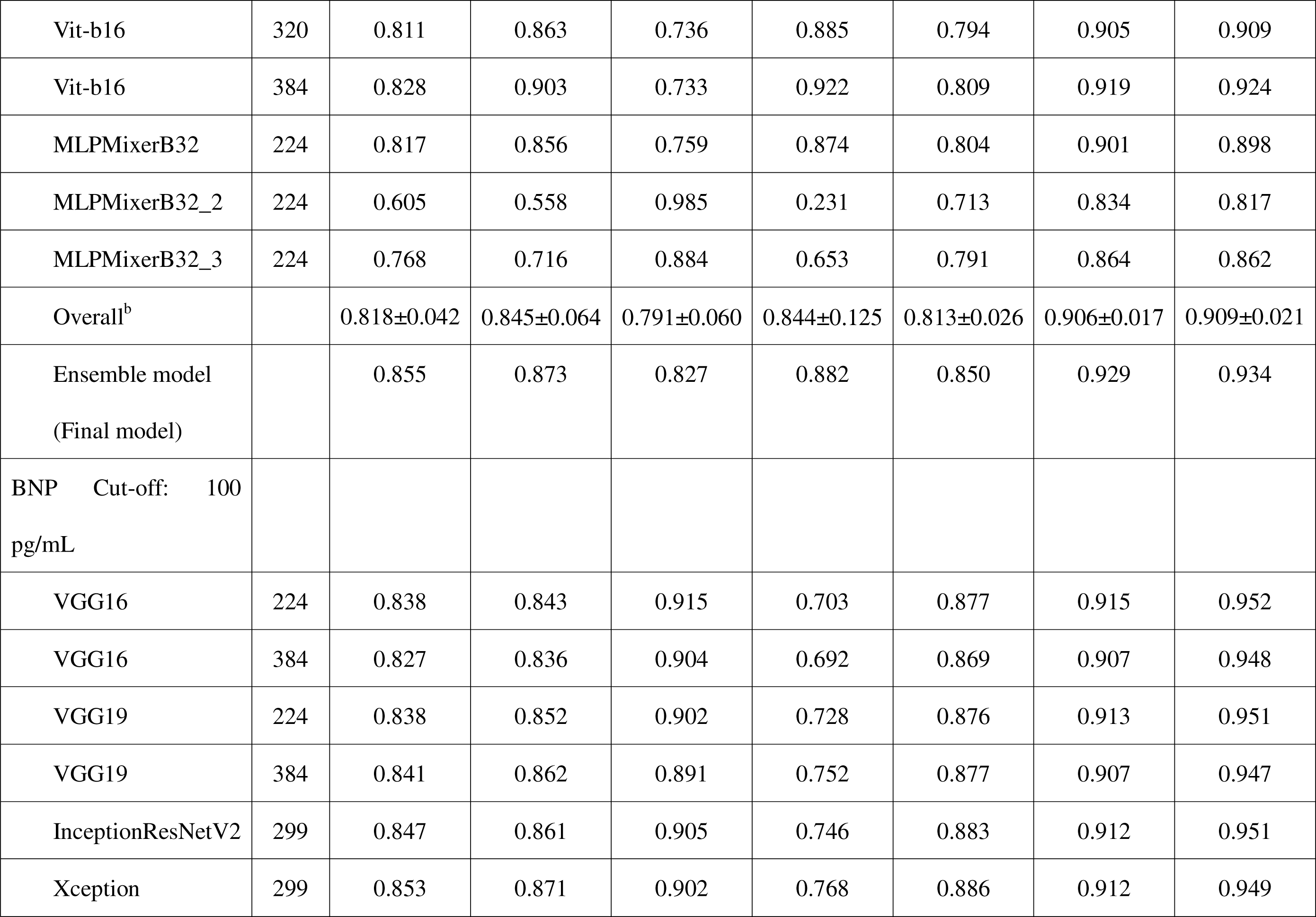
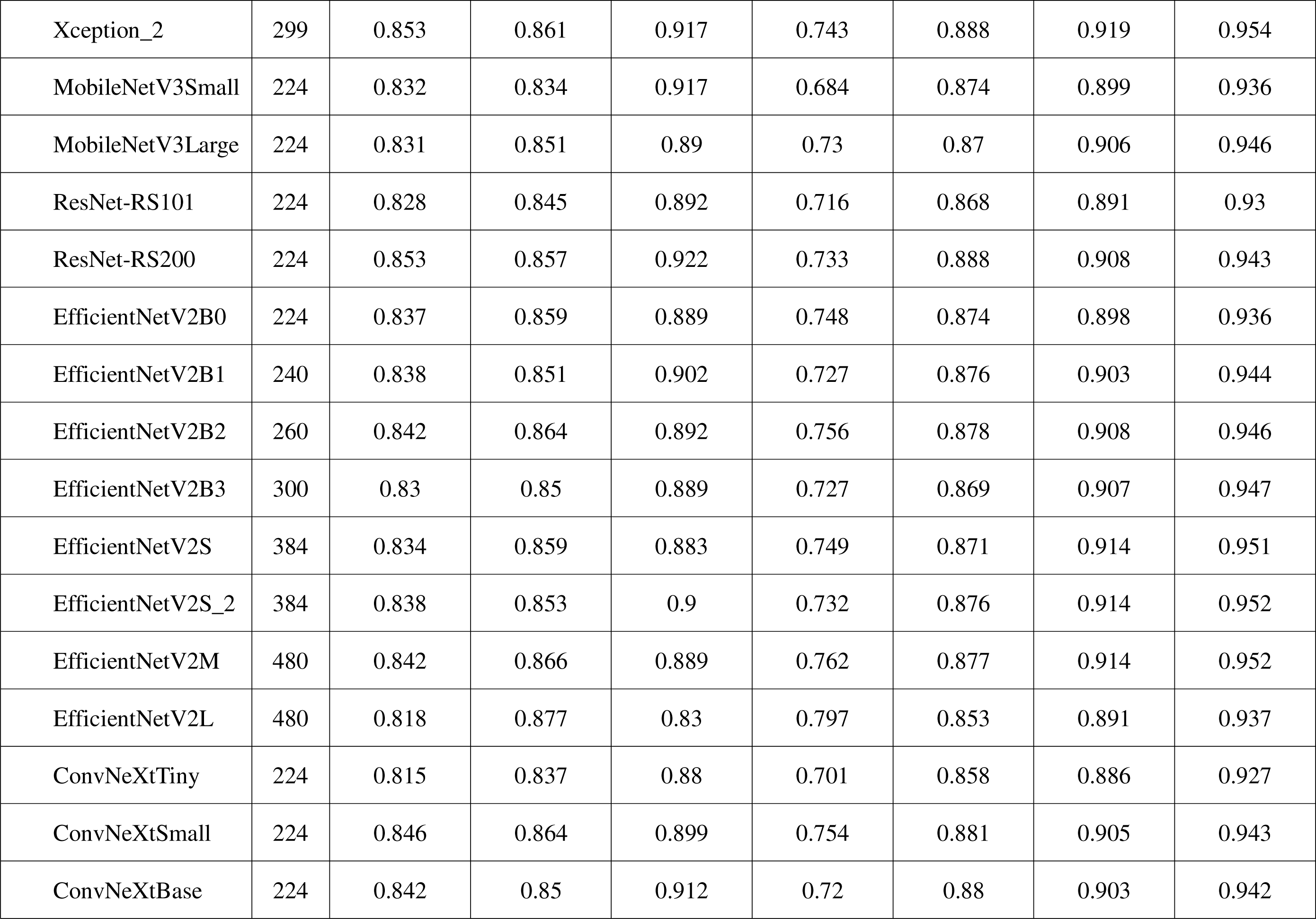
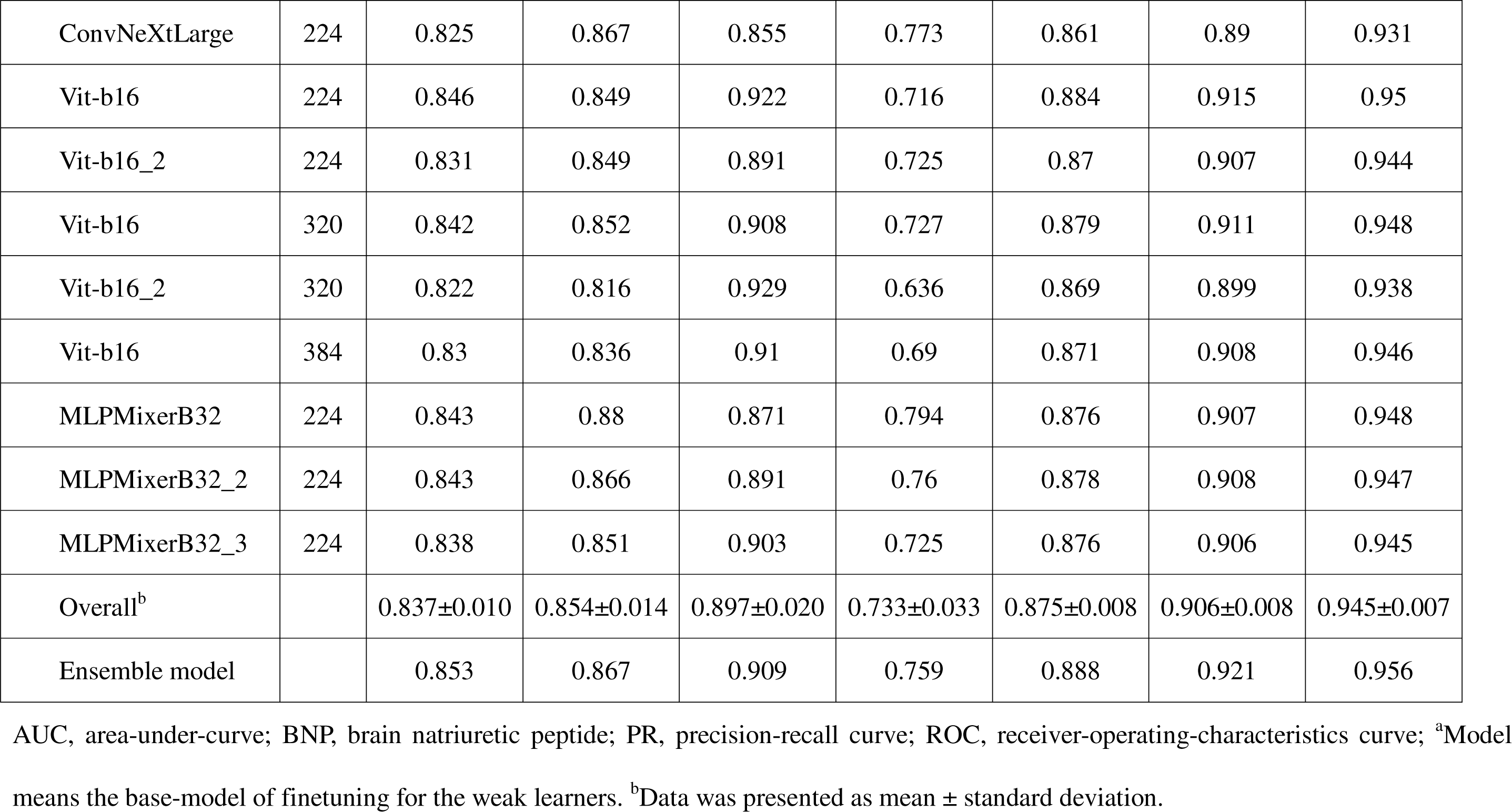
Performance of Models with BNP Cut-Off.

### Performance of Human

A total of 35 participants, including 20 medical doctors of whom 13 were cardiologists, were tested. The duration of medical practice among the participants was 10.2±9.0 years, with 16 identified as experts. The AI model’s performance on the 100 images was as follows: the accuracy 0.920, sensitivity 0.880, specificity 0.957, and f1 score 0.917 (Graphical Abstract). Without the AI assistance, the human participants achieved an accuracy of 0.708±0.049, a sensitivity of 0.693±0.128, and a specificity of 0.722±0.144. With the AI assistance, these measures significantly improved to an accuracy of 0.829±0.068 (P < 0.001), a sensitivity of 0.787±0.113, and a specificity of 0.872±0.097. Even with the AI assistance, no human subjects surpassed the performance of the AI model in terms of accuracy, precision, specificity, or f1 score. The accuracy of the medical doctors and experts was higher than that of non-medical doctors and non-experts in the non-assisted test, respectively (0.725±0.054 vs. 0.687±0.032, P = 0.014; 0.728±0.051 vs. 0.692±0.042, P = 0.030) (Figure 4). However, with the AI assistance, the accuracy of the medical doctors was similar to that of the non-medical doctors (0.818±0.064 vs. 0.843±0.074, P = 0.289), and the accuracy of the non-experts was even higher than that of the experts (0.851±0.074 vs. 0.803±0.054, P = 0.033). In the AI-assisted test, there were 3 non-experts and 1 expert who responded entirely based on the AI model’s predictions, and these four participants achieved the highest accuracy throughout the test, with an accuracy of 0.920. In the non-assisted test, the accuracy had a weak positive correlation with the duration of medical careers (r = 0.414, P = 0.014), while in the AI-assisted test it showed a weak negative correlation (r = −0.347, P = 0.041). For the eight images that were incorrectly predicted by the AI model, the human accuracy was 0.301±0.124. Using majority voting for the hard ensemble prediction, the human accuracy was 0.800 in the initial test and 0.880 with the AI assistance.

**Figure 4.**
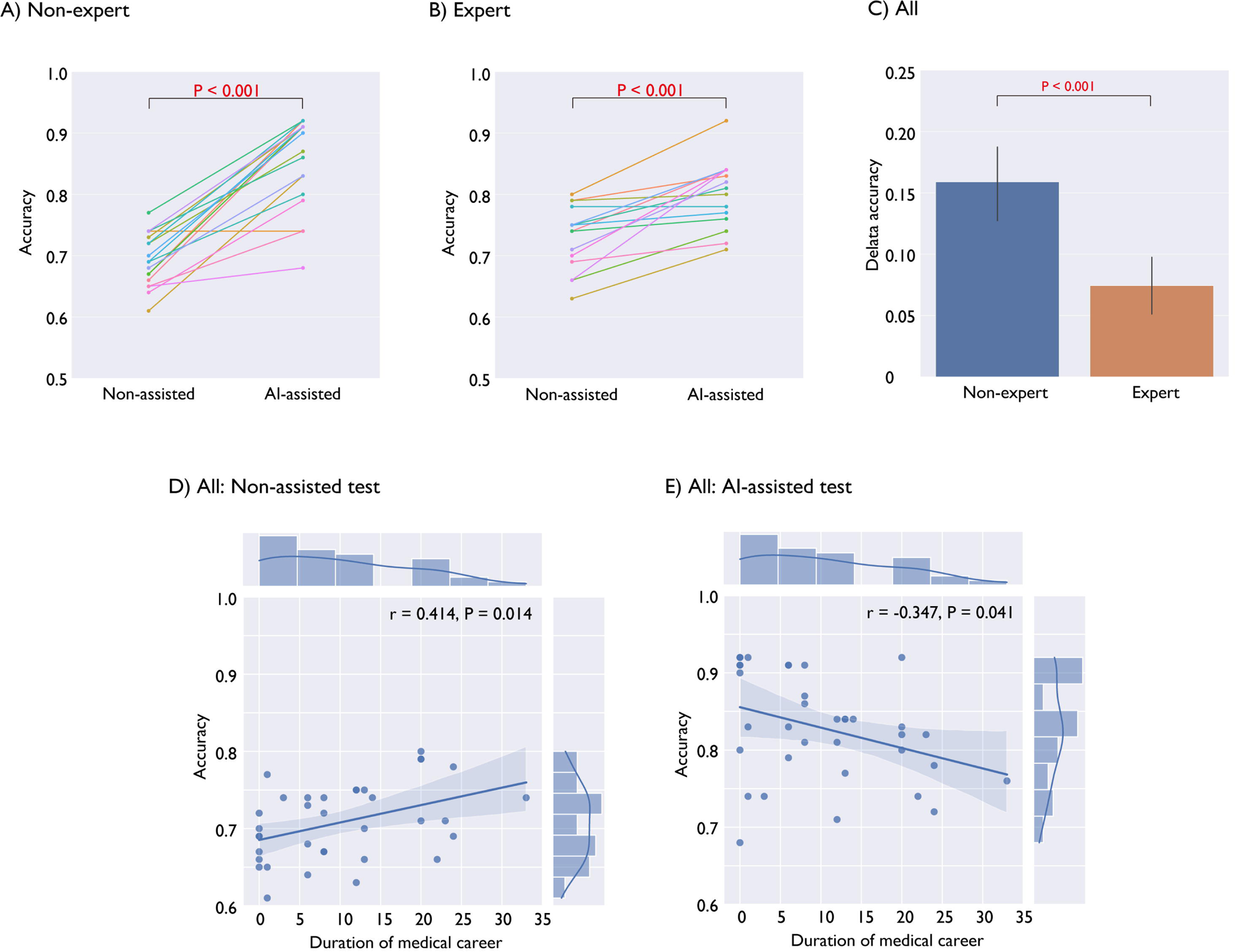
The Results of the Tests. (A and B) Both the non-experts (0.692±0.042 vs. 0.851±0.074, P < 0.001) and experts (0.728±0.051 vs. 0.803±0.054, P < 0.001) improved their accuracy with the assistance of the AI model. (C) The increase in the accuracy with the AI assistance was greater for the non-experts than for the experts (0.159±0.069 vs. 0.074±0.052, P < 0.001). (D) In the non-assisted test, the accuracy had a weak positive correlation with the duration of medical careers (r = 0.414, P = 0.014). (E) However, in the AI-assisted test, it had a weak negative correlation (r = −0.347, P = 0.041). BNP, brain natriuretic peptide; AI, artificial intelligence; AUC, area-under-curve.

### Sub Study

We developed a model to predict BNP values. The mean absolute errors between the predicted and true BNP values were 208 pg/mL, with mean squared errors being 1.01*10^5^ pg^2^/mL^2^. No significant difference was observed between the predicted and true BNP values (P = 0.274). The models’ performances in predicting elevated BNP level using a cut-off of 100 pg/mL was comparable to that of the models using a cut-off of 200 pg/mL (Table 2, Figure 2).

## Discussion

This study presents the development of a high-performing model that predicts elevated BNP levels from chest X-ray images, thereby improving human diagnostic accuracy. Furthermore, this study has revealed the new issue that there exists a gap among participants in the ability to effectively utilize the new AI tool.

The strengths of our study are as follows: First, this is the first report demonstrating that an AI model can predict elevated BNP levels from chest X-ray images, outperforming experienced cardiologists. Our models were predicated on the hypothesis that chest X-ray images contain features associated with elevated BNP levels indicative of heart failure. Prior reports have associated chest X-ray findings such as cardiomegaly, pulmonary venous congestion, interstitial or alveolar oedema, and cephalization with heart failure prediction.^1,3^ These prior evaluations were human based; our study establishes superior diagnostic performance by the AI models in predicting elevated BNP levels, substantiating the one of our hypotheses. The featured map images revealed that our models capture chest X-ray findings akin to prior human reports on chest radiography and heart failure (Figure 1). Nevertheless, various factors such as age, sex, hemoglobin level, renal function, left ventricular end-diastolic pressure, and left ventricular ejection fraction influence plasma BNP levels, so BNP level cannot be perfectly evaluated solely through chest X-ray imaging.^13^ Conversely, the predictability, as indicated by the ROC AUC, was above 0.929 in our test dataset population, suggesting a high level predictability. Matsumoto et al. reported that their model could predict heart failure from chest radiography.^14^ A limitation of their study was that the diagnosis of heart failure was determined by two cardiologists using chest radiographs. Our study highlights the relative inferiority of physician performance in predicting elevated BNP levels compared to the AI model, even among experienced cardiologists. There are several reports predicting pulmonary arterial pressure, pulmonary hypertension, pulmonary wedge pressure, or extravascular lung water from chest radiographs, including similar studies by the same author groups.^15–20^ One limitation of their studies is the difference in timing between when the catheterization was performed and when the X-ray was taken, as the pulmonary artery pressure can greatly vary depending on the patient’s condition such as posture. The performance of the models in the studies is suboptimal, possibly due to the small sample size associated with the invasive catheterization procedures. Zou et al. reported that their model predicted pulmonary hypertension from chest radiographs with an ROC AUC of 0.967; however, in their study, a dataset excluding various conditions, such as pleural/pericardial effusion, and pneumothorax, was utilized. In contrast, our study utilized all available chest radiographs, encompassing a wide range of pathological conditions, which may contribute to the robustness and generalizability of our model.

Second, the performance of our models was benchmarked against front-line physicians. Although numerous AI studies have reported that the AI models could accurately predict medical features from conventional tests, many of these did not evaluate the performance of physicians.

Third, we showed that the AI model’s suggestions could enhance human diagnostic performance, and the utilization gap for new tools is an emerging issue. Regardless of the superior performance of a tool, it is useless if it is not trusted or utilized by users. While many studies have reported that AI models exceed human diagnostic performances, there has yet to be a report detailing the degree of improvement in performance when utilizing AI models, as compared to the standalone performance of the AI models or in the absence of any support. This study discovered that the performance without assistance was positively associated with the duration of medical careers. The AI assistance improved the diagnostic performance of both inexperienced and experienced practitioners. Ironically, the inexperienced ones achieved results comparable to or even surpassing those of the experienced ones. This implies that with the aid of a potent diagnostic tool, inexperienced individuals can perform as well as or even surpass experienced ones. The limited improvement among the experts may be attributed to their confidence in their expertise and skepticism towards the AI model, despite being informed of the AI model’s superior performance compared to any human. Conversely, a less experienced individual might readily accept the AI’s prediction due to lack of confidence. Distrust in new technology or findings and self-confidence will be emerging issues in AI; this kind of skepticism towards novel approaches has always existed in other domains. In particular, the usage of generative AI has begun to be actively discussed. We should strive to understand and adapt appropriately to new ideas, technologies, and tools, including AI.

Fourth, the models used in this study, along with the sample codes for their application, will be made available on GitHub following the publishment of this article. This implies that anyone can evaluate and refine the models, and challenge old notions with new ideas.

Fifth, we enhanced the models using state-of-the-art deep learning techniques used in Kaggle competitions. Our models were based on these technical aspects, and our collected dataset, which was comprehensive and had not been used in previous reports regarding heart failure and chest radiography, is thought to be one of the strengths of our study and may enhance the model’s robustness and generalizability.

Ensemble prediction has the potential to improve performance.^21^ In our study, the accuracy improved from 0.818 in the single model to 0.855 in the final ensemble model. Through the ensemble method, there is a potential enhancement not only in the described performance metrics but also in robustness and generalizability. To enhance the performance of the ensemble, the models’ performances should be reasonably good and their prediction correlation should not be overly strong. This parallels the performance of a heart team, where members who consistently agree with others, or remain silent, contribute little to the quality of decisions, and active members lacking a certain level of performance can impair overall performance. Given that sensitivity and specificity exist in a trade-off relationship, a weak learner that may not be the best in terms of overall accuracy could potentially enhance the accuracy and generalizability of the ensemble model due to the diversity it provides.

Sixth, good old chest radiography is widely available in numerous medical facilities. Technically integrating software, such as the one used in this study, into X-ray machines or smartphones is not challenging. As advancements in both hardware and software continue, the integration process may become even more streamlined, potentially allowing these tools to be used in diverse ways, with the potential to change the world.

### Study Limitations

Natriuretic peptides are used for diagnosis of heart failure, employing either absolute values or relative changes, and for managing the condition through sequential relative changes. This study primarily conducted with binary prediction, as the main goal was to diagnose heart failure in this time. Notably, the human performance in predicting absolute BNP values from chest X-ray images was extremely poor in preliminary testing (data not shown). The choice of a cut-off BNP value warrants discussion. The cut-off values should be determined based on intended purpose while ensuring a balance between sensitivity and specificity. We set the BNP cut-off value at 200 pg/mL with the aim of diagnosing unrecognized heart failure that requires early management; however, a cut-off value of 100 or 125 pg/mL would be considered for different purposes or applications. In this study, models with a cut-off of 100 pg/mL demonstrated performance comparable to those with a cut-off of 200 pg/mL. While we labelled the X-ray images based on the BNP cut-off value, it is important to remember that heart failure is not diagnosed based solely on natriuretic peptide values. One of our goals is to diagnose unrecognized heart failure that needs early intervention, which is not synonymous with diagnosing elevated BNP levels alone.

## Conclusions

The AI model can predict elevated BNP levels from chest X-ray images with superior performance compared to experienced cardiologists and can improve the diagnostic performance of individuals, ranging from non-experts to experienced cardiologists. The gap in utilizing new tools represents one of the emerging issues.

## Data Availability

The source codes and weights of the models used in this study will be available on GitHub.

https://github.com/ekagawa007/kagawa_for_review

## Acknowledgements

We express our gratitude to all the participants who participated in this study.

