## Supplementary material for "Predicting elevated natriuretic peptide in chest radiography: Emerging utilization gap for artificial intelligence": Supplemantary Materials

**Table of Contents:**

**Section S1: Supplemental Methods**

**Section S2: Supplemental Table**

**Section S3: References for the Supplemental Appendix**

### Section S1: Supplemental Methods

The patients who were duplicated in both hospitals were assigned to only one dataset. Patients taking an angiotensin receptor neprilysin inhibitor were included because we intended to make models robust and widely available in real clinical settings, and it might not increase BNP.[1] The models were trained using an NVIDIA GeForce RTX 3080Ti GPU (NVIDIA Corporation, Santa Clara, CA, USA) or the Google Colaboratory platform (Google LLC, Mountain View, CA, USA).

#### Image Pre-processing

Non-square chest X-ray images were transformed to square using 0 padding (black field), not cropping. We planned to pre-process the images in which the head was in the upper half of the image and the spine was at 60 ° - 120 ° angle to the horizontal line of images. However, all the images met these requirements, so these image pre-processing steps were not utilized.

#### Training of Models (Weak Learners)

The summary of the fine-tuning models, which were based on well-known image recognition models, is shown on GitHub. These models were loaded with weights that had been pre-trained by ImageNet, a large non-medical image dataset.[2] The top layers of the fine-tuned models were designed as two full connected layers with 2048 and 2 neurons, respectively, each layer having a preceding dropout layer. The provability of the BNP >= 200 pg/mL was determined by two neurons with softmax activation function in the final output layers, generating numbers ranging between 0 and 1. Data augmentation with random width shifts up to 10%, height shifts up to

15%, rotation up to 15 °, horizontal flips, Mixup, RandomErasing, and CutMix techniques were utilized during model training.[3-5] We applied Label Smoothing to regularize the neural networks.[6] All models in this study were optimized using sharpness aware minimization with RAdam or Adam.[7,8] The hyperparameters are listed in Supplemental Table 2. The input image sizes of the models were determined based on the pre-training image sizes. The training process was visualized with the accuracy and loss of the training and validation datasets for every epoch, with all models trained for at least 100 epochs. If overfitting was observed or learning seemed to have converged by 100 epochs, training was terminated. If learning had not converged at 100 epochs, training was extended for an additional 50 epochs until overfitting or learning convergence was confirmed. The weights of the models that achieved the minimum loss in the validation dataset at the end of each epoch were saved and used for the study. These models served as candidate for weak learners for the final ensemble model.

**Sub Study**

We developed models to predict the BNP values. Images with BNP values >= 2000 pg/mL were excluded from the three datasets; the top two full connected layers were replaced with three full connected layers (two layers of 1024 neurons with activation function mish and one layer with one neuron with sigmoid function). We trained the models using the last 11 layers, which are trainable for fine-tuning. Additionally, we trained models to predict elevated BNP values with a cut-off of 100 pg/mL, modifying hyperparameters based on the training from the main study.

**Section S2: Supplemental Table**

**Supplemental Table 1. The Hyperparameters of the Models**

| Model | Optimizer | Batch size | Epoch | Trainable Layer | Initial learning rate | Minimum learning rate |
|---|---|---|---|---|---|---|
| VGG16 | SAM, RAdam | 32 | 1 – 10 | Top 3 | 0.001 | CA alpha 0.01 |
| | | | 11 – 30 | Top 10 | 0.0001 | CA alpha 0.01 |
| | | | 31 – | All | 0.00001 | CA alpha 0.01 |
| VGG19 | SAM, RAdam | 16 | 1 – 10 | Top 3 | 0.001 | CA alpha 0.01 |
| | | | 11 – 30 | Top 10 | 0.0001 | CA alpha 0.01 |
| | | | 31 – | All | 0.00001 | CA alpha 0.01 |
| InceptionResNetV2 | SAM, RAdam | 16 | 1 – 10 | Top 3 | 0.001 | CA alpha 0.01 |
| | | | 11 – 30 | Top 155 | 0.0001 | CA alpha 0.01 |
| | | | 31 – | All | 0.0001 | CA alpha 0.01 |
| Xception | SAM, RAdam | 8 | 1 – 10 | Top 3 | 0.001 | CA alpha 0.01 |
| | | | 11 – 30 | Top 31 | 0.0001 | CA alpha 0.01 |
| | | | 31 – | All | 0.0001 | CA alpha 0.01 |
| MobileNetV3Small | SAM, RAdam | 16 | 1 – 10 | Top 3 | 0.001 | CA alpha 0.01 |
| | | | 11 – 30 | Top 34 | 0.0001 | CA alpha 0.01 |
| | | | 31 – | All | 0.0001 | CA alpha 0.01 |
| MobileNetV3Large | SAM, RAdam | 8 | 1 – 10 | Top 3 | 0.001 | CA alpha 0.01 |
| | | | 11 – 30 | Top 74 | 0.0001 | CA alpha 0.01 |
| | | | 31 – | All | 0.0001 | CA alpha 0.01 |
| ResNet-RS101 | SAM, | 256 | 1 – 10 | Top 3 | 0.001 | CA alpha 0.01 |

| | | | | | | |
|---|---|---|---|---|---|---|
| | RAdam | | | | | |
| | | | 11 – 30 | Top 50 | 0.0001 | CA alpha 0.01 |
| | | | 31 – | All | 0.0001 | CA alpha 0.01 |
| ResNet-RS200 | SAM, RAdam | 32 | 1 – 10 | Top 3 | 0.001 | CA alpha 0.01 |
| | | | 11 – 30 | Top 57 | 0.0001 | CA alpha 0.01 |
| | | | 31 – | All | 0.0001 | CA alpha 0.01 |
| EfficientNetV2B0 | SAM, RAdam | 64 | 1 – 10 | Top 3 | 0.001 | CA alpha 0.01 |
| | | | 11 – 30 | Top 126 | 0.0001 | CA alpha 0.01 |
| | | | 31 – | All | 0.0001 | CA alpha 0.01 |
| EfficientNetV2B1 | SAM, RAdam | 32 | 1 – 10 | Top 3 | 0.001 | CA alpha 0.01 |
| | | | 11 – 30 | Top 141 | 0.0001 | CA alpha 0.01 |
| | | | 31 – | All | 0.0001 | CA alpha 0.01 |
| EfficientNetV2B2 | SAM, RAdam | 16 | 1 – 10 | Top 3 | 0.001 | CA alpha 0.01 |
| | | | 11 – 30 | Top 156 | 0.0001 | CA alpha 0.01 |
| | | | 31 – | All | 0.0001 | CA alpha 0.01 |
| EfficientNetV2B3 | SAM, RAdam | 8 | 1 – 10 | Top 3 | 0.001 | CA alpha 0.01 |
| | | | 11 – 30 | Top 175 | 0.0001 | CA alpha 0.01 |
| | | | 31 – | All | 0.0001 | CA alpha 0.01 |
| EfficientNetV2S | SAM, RAdam | 8 | 1 – 10 | Top 3 | 0.001 | CA alpha 0.01 |
| | | | 11 – 30 | Top 217 | 0.0001 | CA alpha 0.01 |
| | | | 31 – | All | 0.0001 | CA alpha 0.01 |
| EfficientNetV2M | SAM, RAdam | 4 | 1 – 10 | Top 3 | 0.001 | CA alpha 0.01 |
| | | | 11 – 30 | Top 77 | 0.0001 | CA alpha 0.01 |
| | | | 31 – | All | 0.0001 | CA alpha 0.01 |

| | | | | | | |
|---|---|---|---|---|---|---|
| EfficientNetV2L | SAM, RAdam | 2 | 1 – 10 | Top 3 | 0.001 | CA alpha 0.01 |
| | | | 11 – 30 | Top 107 | 0.0001 | CA alpha 0.01 |
| | | | 31 – | All | 0.0001 | CA alpha 0.01 |
| ConvNeXtTiny | SAM, RAdam | 16 | 1 – 10 | Top 3 | 0.001 | CA alpha 0.01 |
| | | | 11 – 30 | Top 31 | 0.0001 | CA alpha 0.01 |
| | | | 31 – | All | 0.0001 | CA alpha 0.01 |
| ConvNeXtSmall | SAM, RAdam | 8 | 1 – 10 | Top 3 | 0.001 | CA alpha 0.01 |
| | | | 11 – 30 | Top 31 | 0.0001 | CA alpha 0.01 |
| | | | 31 – | All | 0.0001 | CA alpha 0.01 |
| ConvNeXtBase | SAM, RAdam | 8 | 1 – 10 | Top 3 | 0.001 | CA alpha 0.01 |
| | | | 11 – 30 | Top 31 | 0.0001 | CA alpha 0.01 |
| | | | 31 – | All | 0.0001 | CA alpha 0.01 |
| ConvNeXtLarge | SAM, RAdam | 2 | 1 – 10 | Top 3 | 0.001 | CA alpha 0.01 |
| | | | 11 – 30 | Top 31 | 0.0001 | CA alpha 0.01 |
| | | | 31 – | All | 0.0001 | CA alpha 0.01 |
| ConvNeXtXLarge | SAM, RAdam | 2 | 1 – 10 | Top 3 | 0.001 | CA alpha 0.01 |
| | | | 11 – 30 | Top 31 | 0.0001 | CA alpha 0.01 |
| | | 1 | 31 – | Top 167 | 0.0001 | CA alpha 0.01 |
| Vit-b16 | SAM, RAdam | 8 | 1 – 10 | Top 3 | 0.001 | CA alpha 0.01 |
| | | | 11 – 30 | Top 11 | 0.0001 | CA alpha 0.01 |
| | | | 31 – 100 | All | 0.0001 | CA alpha 0.01 |
| MLPMixerB32 | SAM, RAdam | 8 | 1 – 10 | Top 3 | 0.001 | CA alpha 0.01 |
| | | | 11 – 30 | Top 30 | 0.0001 | CA alpha 0.01 |

| | | | | | | |
|---|---|---|---|---|---|---|
| | | | 31 – 100 | All | 0.0001 | CA alpha 0.01 |

Activation function of the second to last full connected layer is mish. CA indicates cosine annealing; LS, label smoothing; RAdam, Rectified Adam; SAM, sharpness aware minimization.
